# The COVID-19 Healthcare Personnel Study (CHPS): Overview, Methods and Preliminary Findings

**DOI:** 10.1101/2020.10.29.20222372

**Authors:** Charles DiMaggio, David Abramson, Ezra S. Susser, Christina W. Hoven, Qixuan Chen, Howard F Andrews, Daniel Herman, Jonah Kreniske, Megan Ryan, Ida Susser, Lorna E. Thorpe, Guohua Li

**Affiliations:** Department of Surgery, NYU School of Medicine; NYU Global Institute for Public Health, New York University; Mailman School of Public Health, Columbia University; New York State Psychiatric Institute, Mailman School of Public Health, Columbia University; Department of Biostatistics, Mailman School of Public Health, Columbia University; Data Coordinating Center (DCC), Columbia University Irving Medical Center, NY State Psychiatric Institute; Silberman School of Social Work, Hunter College, CUNY; Department of Global Health and Social Medicine, Harvard Medical School; Global Psychiatric Epidemiology Group, New York State Psychiatric Institute; Department of Anthropology, Hunter College and Graduate Center, City University of New York; Department of Population Health, NYU Grossman School of Medicine; Department of Anesthesiology, Columbia University Medical Center

## Abstract

**Introduction:** The onslaught of the COVID-19 pandemic has placed severe demands on US health systems and the health care workforce. In New York State (NYS) and New York City (NYC), the early American epicenter, hospitals ran the risk of exhausting supplies of ventilators, ICU beds, and personal protective equipment (PPE); the capacity of the healthcare workforce to meet the added demand was equally strained. The COVID-19 Healthcare Personnel Study (CHPS) was designed to assess adverse short and long-term physical and mental health impacts of the COVID-19 pandemic on New York’s physicians, nurse practitioners, and physician assistants.

**Methods:** Population-based online survey of physicians, nurse practitioners and physician assistants registered with the New York State Department of Health. Survey-weighted descriptive results were analyzed using frequencies, proportions, and means, with 95% confidence intervals. Odds ratios were calculated for association using survey-weighted logistic regression.

**Results:** Over half (51.5%; 95% CI 49.1, 54.0) of respondents worked directly with COVID-19 patients. Among those tested, 27.3% (95% CI 22.5, 32.2) were positive. The majority (57.6%; 95% CI 55.2, 60.0) of respondents reported that the COVID-19 pandemic had a negative impact on their mental health. Negative mental health was associated with experiencing symptoms of COVID-19 (OR=1.7, 95% CI 1.3, 2.1) and redeployment to unfamiliar functions. (OR=1.3, 95% CI 1.1, 1.6).

**Conclusions:** A majority of New York healthcare providers were involved in treating COVID-19 patients and reported that the pandemic had a negative impact on their mental health.

## Introduction

The sudden onslaught of the COVID-19 pandemic in spring 2020 placed severe demands on US health systems and the health care workforce. Hospitals ran the risk of exhausting their supplies of ventilators, ICU beds, and personal protective equipment (PPE); the capacity of the health care workforce to meet the added demand was equally strained. In the late winter and early spring of 2020, New York State (NYS) and New York City bore a disproportional share of the burden, with approximately half of all confirmed cases in America. NYS instituted a number of strategies to expand hospital capacity and the workforce: the governor mandated that all hospitals increase bed capacity by 50%; specialized hospital facilities were constructed in large convention spaces; efforts to purchase and obtain donated PPE were accelerated; volunteer, retired, and student health care professionals were enlisted to supplement the workforce; and hospitals and health systems explored ways of repurposing and expanding their stock of critical equipment.

The COVID-19 Healthcare Personnel Study (CHPS) was launched to longitudinally assess the adverse health impacts of the COVID-19 pandemic on New York State (NYS) health professionals. Between 28 April and 30 June 2020, in the immediate weeks after the pandemic crested in NYS, CHPS collaborators recruited members of multiple health care professions across NYS to participate. We present here a description of the methods and the initial results of CHPS, based on the first wave of responses from physicians, nurse practitioners and physician assistants. We report key characteristics of respondents, their exposure to and experience with treating COVID-19 patients, and the impact of the pandemic on their practice patterns and health.

## Methods

On 28 April 2020, an invitation to participate in the CHPS survey was sent from the office of the NYS Commissioner of Health to 103,103 physicians, 21,503 nurse practitioners and 14,503 physician assistants, representing all providers among those professions licensed to practice in the state, for a total of 139,109 emails. This baseline survey was closed to enrollment on 30 June 2020.

The survey was administered through REDCap, a secure web application managed by Data Coordinating Center at the New York State Psychiatric Institute (NYSPI) for building and managing online surveys and databases.^1^ Data were downloaded and read into the R statistical computing system,^2^ checked for outliers and cleaned. We used raking procedure to assign a survey weight for each respondent to make the sample representative of the target population of physicians, nurse practitioners and medical assistants in age, gender and geographic location across 10 regions of New York State.^3, 4, 5, 6, 7, 8^.

Statistical analyses consisted of survey-weighted counts, proportions, and means with 95% confidence intervals using the R “survey” package.^9^ Odds ratios for association were calculated using survey-weighted logistic regression models. The protocol was approved by the Institutional Review Boards (IRB) of Columbia University Medical Center, the New York State Psychiatric Institute, the City University of New York, and NYU School of Medicine. The questionnaire is available on request to the CHPS Steering Committee.

## Results

Of the 139,109 emails sent, 38,090 (27.4%) reached an intended recipient. Of these, 2,076 physicians, nurse practitioners and physician assistants completed the survey, for a response rate of 5.5%. The sample had a weighted mean age of 50.4 (95% CI 49.9, 51.0), of whom 46.1% (95% CI 43.7, 48.5) were female. 68.7% (95% CI 66.6, 70.9) of the weighted sample usually practiced in the New York City Metro area (5 boroughs of New York City, Nassau, Suffolk, Rockland, and Westchester Counties), and 67.7% (95% CI 65.5, 69.9) were practicing in the NYC Metro area at the time they completed the survey. 30% (95% CI 30.18, 30.22) practiced in primary care; 46.5% (95% CI 44.1, 49.0) of respondents practiced primarily in a hospital setting.

51.5% (95% CI 49.1, 54.0) or an estimated frequency of 69,586 (95% CI 65,572, 73,599) NYS physicians, nurse practitioners and physician assistants reported having worked directly or in close physical contact with COVID-19 patients. Nearly a quarter of all respondents (24.1%; 95% CI 21.6, 26.5) reported changing their living arrangements because of concern about exposure to COVID-19. An estimated frequency of 1,159 (95% CI 586, 1,733) NYS MD/NP/PA’s came out of retirement to work on the COVID-19 response. One third (32.8%, 95% CI 30.5, 35.1) of respondents indicated that they were required to perform functions different than their usual practice in response to COVID-19.

A large proportion of respondents (43.3%; 95% CI 40.9, 45.8) reported reluctance to work directly with COVID-19 patients, primarily because of fear of infecting oneself or others. (Table 1) Over a third the weighted sample (37.7%, 95% CI = 35.3, 40.0) reported shortages of N95 masks, and over half (51.5%, 95% CI= 48.6, 54.1) reported having had to re-use disposable personal protective equipment in a manner which seemed unsafe. 22.8% (20.7, 25.0) reported shortages of test kits, and nearly a fifth (19.5%, 95% CI = 17.6, 22.0) reported shortages of personnel. A smaller proportion reported shortages of ventilators (2%, 95% CI = 1.4, 3.0) or beds (4.4%, 95% CI = 3.4, 6.0).

**Table 1.** Weighted Percent (95% Confidence Interval) of Reasons for Reluctance in Treating Covid-19 Patients Among Those Expressing Reluctance. New York State COVID-19 Health Provider Survey. First Wave. Physicians, Nurse Practitioners, Physician Assistants. 28 April – 30 June 2020.

|  | Unweighted<br>Frequency<sup>1</sup> | Weighted<br>Frequency<sup>2</sup> | Weighted<br>Percent (95% CI) |
| --- | --- | --- | --- |
| Fear of Infecting Self | 696 | 46,297 | 34.1 (31.8, 36.0) |
| Fear of Infecting Others | 675 | 44,292 | 32.6 (30.4, 35.0) |
| Pre-Existing Health Condition | 322 | 20,737 | 15.3 (13.6, 17.0) |
| Insufficient Protective Equipment | 400 | 26,876 | 19.8 (17.9, 22.0) |
| Insufficient Skills or Expertise | 276 | 16,710 | 12.3 (10.9, 14.0) |

<sup>1</sup> Total Unweighted Frequency 2,076
<sup>2</sup> Total Weighted Frequency 135,722

28.0% (95% CI 25.7, 30.3) of the weighted sample attempted to obtain a test for COVID-19. Of these, 21.4% (95% CI 19,4, 23.5) were able to obtain a test; 27.3% (95% CI 22.5, 32.2) of these tests were positive.

Well over half of the respondents (57.6%) reported that the COVID-19 pandemic had a negative impact on their mental health, and 43% felt it had negatively impacted personal relationships. (Table 2) Respondents who indicated that they were redeployed or required to perform different functions in response to COVID-19 were more likely to report negative mental health impacts (OR=1.3, 95% CI 1.1, 1.6) Reporting symptoms of COVID-19 was also associated with adverse mental health (OR=1.7, 95% CI 1.3, 2.1).

**Table 2.** Weighted Percent (95 % Confidence Interval) of Respondents Experiencing Negative Impacts as a Result of COVID-19. New York State COVID-19 Health Provider Survey. First Wave. Physicians, Nurse Practitioners, Physician Assistants. 28 April – 30 June 2020.

|  | **Unweighted**<br>**Frequency<sup>1</sup>** | **Weighted**<br>**Frequency<sup>2</sup>** | **Percent**<br>**(95% CI)** |
| --- | --- | --- | --- |
| Mental Health | 1196 | 78201 | 57.6 (55.2, 60.0) |
| Physical Health | 656 | 44126 | 32.5 (30.0, 35.0) |
| Ability to work | 741 | 50046 | 36.9 (34.5, 39.0) |
| Relationships | 915 | 59531 | 43.9 (41.4, 46.0) |

<sup>1</sup> Total Unweighted Frequency 2,076
<sup>2</sup> Total Weighted Frequency 135,722

## Discussion

This study adds to early reports from China that “a considerable proportion of health care workers (treating COVID-19 patients) reported experiencing symptoms of depression, anxiety, insomnia, and distress”, ^10^ as well as a survey of Saudi HCWs who responded to the 2014 MERS outbreak in which “almost two thirds reported having psychological problems”^11^. Our results indicate that mental distress was associated with redeployment to non-familiar clinical roles.

The literature on the mental health effects of COVID-19 on HCWs remains nascent, but we can look to research on the effect SARS in the early 2000s and more recent MERS-CoV outbreaks as a guide. A 2003 survey of 769 Canadian HCWs found “higher levels of burnout (p = 0.019), psychological distress (p<0.001), and posttraumatic stress (p<0.001)” among those who had treated SARS patients compared to those who did not, one to two years after the outbreak.^12^ In an overview of the experience of HCWs responding to the Toronto SARS outbreak in 2003, the investigators “estimated that a high degree of distress was experienced by 29-35% of hospital workers.” They concluded there were “Three categories of contributory factors…being a nurse, having contact with SARS patients and having children.”^13^

There is evidence supporting specific measures to address distress among HCWs responding to pandemics. Among helpful factors reported by HCWs in coping with a MERS-CoV outbreak that occurred in Jeddah, Saudi Arabia were a “Positive attitude from colleagues in your department” and financial compensation.**Error! Bookmark not defined**. The authors of a series of studies of HCW attitudes toward responding to SARS concluded that “Reducing pandemic-related stress may best be accomplished through interventions designed to enhance resilience in psychologically healthy people,” by providing psychological first aid, institutional training, support and leadership.^14^ The same authors also pointed to the importance of practical and tangible support,^13^ as well as the potential utility of computer-based tools to improve confidence and self-efficacy.^15^

Our data also support a high risk of contracting COVID-19 among HCWs carrying out their professional responsibilities, with 27% of clinicians who were able to access a test for COVID-19 testing positive. Important and meaningful challenges like a lack of personal protective equipment may contribute to that risk,^16^ though it is important to note that during a similar time period tests for SARS CoV-2 in NYS were in short supply^17^ and often reserved for the sickest patients, and the overall NYS positivity rate was approximately 40%.^18^

Our study is subject to a number of important limitations. These results apply narrowly to physicians and advanced practice clinicians. The demographics of this group differ from other groups demonstrated to be at increased risk of COVID-19, including Blacks/African Americans who make up a large proportion of persons providing non-medical direct services to COVID-19 patients in NYC, but may not be well represented among physicians and advanced practice clinicians.^19^ This is an area in which additional results from the CHPS addressing nursing and non-medical providers can provide insights.

Our response rate was low, in part because the survey was released to the study population during the height of the Pandemic in NY state. Hospitalizations were still high, as was the workload of responding health professionals(including those in our study). Given these taxing external circumstances, health professionals may not have had the necessary time/energy to complete a voluntary survey. Despite these impediments, our response rate was comparable with other recent studies of physicians.^20^ Still, while we utilized statistical procedures to align our survey-adjusted sample with important demographics of the target population, **Error! Bookmark not defined**.,**Error! Bookmark not defined**.,**Error! Bookmark not defined**.,**Error! Bookmark not defined**.,**Error! Bookmark not defined**. our sample may be biased.

We conclude that at the height of the COVID-19 pandemic in New York State in early 2020, healthcare workers, including the majority of physicians, nurse practitioners and physician assistants, experienced work-related stressors (including an increased risk of COVID-19 exposure, redeployment and resource shortages), which rendered them vulnerable to impairments in both their mental/physical health, as well as their personal relationships. Given that the COVID-19 Pandemic is an ongoing health crisis, potential interventions to help lessen the work-related stressors of these health professionals(e.g. preventing resource shortages, redeployment training/support, etc.) are needed to mitigate these types of impairments in the future.

## Data Availability

Researchers may contact study principal investigators to request access to data.

## About the COVID-19 Healthcare Personnel Study (CHPS)

CHPS is a collaborative enterprise representing a concerted effort by health researchers from multiple public and private New York academic institutions, including the Hunter College-City University of New York, Columbia University, the New York State Psychiatric Institute (NYSPI), New York University (NYU), and NYU Langone Health. The primary contacts for the study are Drs. Guohua Li (Columbia University), Charles DiMaggio (NYU Langone), and David Abramson (NYU). The Steering Committee consists of the primary contacts and Drs. Ezra Susser and Christina Hoven (Columbia University-NYSPI). Co-Investigators include Drs. Lorna Thorpe (NYU Langone); Howard Andrews (NYSPI); Dan Herman and Elizabeth Cohn (Hunter College); and Jonah Kreniske (Research Fellow Harvard Medical School).

## Acknowledgments

We thank Barbara Lang for administrative support, Dr. Jennifer Norton for data cleaning and preparation, and Dr. Howard Zucker and the office of the New York State Commissioner of Health for inviting the NYS Healthcare workforce to participate in this survey.

## Citations

1 REDCap. https://www.project-redcap.org/. Accessed 28 July 2020

2 R Core Team (2019). R: A language and environment for statistical computing. R Foundation for Statistical Computing, Vienna, Austria. URL https://www.R-project.org/.

3 Lumley (2004) Analysis of complex survey samples. Journal of Statistical Software 9(1):1-19

4 State of New York. Metrics to Guide Reopening New York. https://forward.ny.gov/metrics-guide-reopening-new-york Accessed 28 July 2020

5 New York Physician Workforce Profile 2014 Edition, The New York Health Workforce Data System, School of Public Health, University at Albany, State University of New York. https://www.chwsny.org/our-work/reports-briefs/r-2015-6/. Accessed 15 July 2020

6 Martiniano R, Wang S, Moore J. A Profile of New York State Nurse Practitioners, 2017. Rensselaer, NY: Center for Health Workforce Studies, School of Public Health, SUNY Albany; October 2017.

7 Liu Y, Martiniano R, Moore J. A Comparative Analysis of New York’s Active Nurse Practitioners and Physician Assistants. Rensselaer, NY: Center for Health Workforce Studies, School of Public Health, SUNY Albany; May 2018.

8 State of New York Deptartment of Education. PA Licenses by County. http://www.op.nysed.gov/prof/med/medcounts.htm. Accessed 28 July 2020

9 Lumley T. Analysis of complex survey samples. Journal of Statistical Software 9(1): 1–19. (2004)

10 Lai, J, Ma, S, Wang, Y, et al. Factors Associated With Mental Health Outcomes Among Health Care Workers Exposed to Coronavirus Disease 2019. JAMA Network Open, 3(3), e203976–e203976. doi:10.1001/jamanetworkopen.2020.3976

11 Alsahafi, A.J. and Cheng, A.C. Knowledge, Attitudes and Behaviours of Healthcare Workers in the Kingdom of Saudi Arabia to MERS Coronavirus and Other Emerging Infectious Diseases. Int. J. Environ. Res. Public Health 2016, 13, 1214.

12 Maunder RG, Lancee, WJ, Balderson, KE, et al. Long-term Psychological and Occupational Effects of Providing Hospital Healthcare during SARS Outbreak. Emerg Infect Dis. Volume 12, Number 12—December 2006

13 Maunder, R. The experience of the 2003 SARS outbreak as a traumatic stress among frontline healthcare workers in Toronto: lessons learned. Philosophical Transactions of the Royal Society B. Biological Sciences. 29 July 2004 Volume 359 Issue 1447.

14 Maunder, RG, Leszcz M, Savage D, et al. Applying the Lessons of SARS to Pandemic Influenza: An Evidence-based Approach to Mitigating the Stress Experienced by Healthcare Workers. Canadian Journal of Public Health volume 99, pages 486–488 (2008).

15 Maunder RG, Lancee WJ, Mae R, et al. Computer-assisted resilience training to prepare healthcare workers for pandemic influenza: a randomized trial of the optimal dose of training. BMC Health Serv Res. 10, Article number: 72 (2010)

16 Nguyen LH, Drew DA, Graham MS et al. Risk of COVID-19 among front-line health-care workers and the general community: a prospective cohort study. Lancet. 31 July 2020. 5(9):E475–E483. https://doi.org/10.1016/S2468-2667(20)30164-X (Accessed 18 September 2020)

17 Feuer W. New York City doctor says he has to ‘plead to test people’ for coronavirus. CNBC. Health and Science. 2 March 2020. https://www.cnbc.com/2020/03/02/coronavirus-new-york-city-doctor-has-to-plead-to-test-people.html Accessed 15 October 2020.

18 New York State. Percentage Positive Results By Region Dashboard. https://forward.ny.gov/percentage-positive-results-region-dashboard Accessed 18 September 2020.

19 Hong N. 3 hospital workers gave out masks. Weeks later, they all were dead. New York Times. Published online May 2020. https://www.nytimes.com/2020/05/04/nyregion/coronavirus-ny-hospital-workers.html Accessed 9 May 2020

20 West CP, Dyrbye LN, Sinsky C, et al. Resilience and Burnout Among Physicians and the General US Working Population. JAMA Netw Open. 2020;3(7):e209385. doi:10.1001/jamanetworkopen.2020.9385. Accessed 14 October 2020

